# Scalability of mobile technology interventions in the prevention and management of HIV among adolescents in low- and middle-income countries: protocol for a systematic review

**DOI:** 10.1101/2023.03.21.23287533

**Authors:** Emmanuel Adebayo, Dongqing Wang, Adesola O. Olumide, Adesola Ogunniyi, Wafaie Fawzi

**Affiliations:** Adolescent Health Unit, Institute of Child Health, College of Medicine, University of Ibadan, Ibadan, Nigeria; Department of Global and Community Health, College of Health and Human Services, George Mason University, Virginia, USA; University College Hospital, Ibadan, Nigeria; Department of Medicine, College of Medicine, University of Ibadan, Ibadan, Nigeria; Department of Global Health and Population, Harvard T.H. Chan School of Public Health, Boston, USA

**Keywords:** HIV Prevention and Management, Adolescents, mobile technology, scalable interventions

## Abstract

**Introduction:** The rate of new infection of HIV is still high among adolescents globally. Adolescents in low and middle-income countries (LMICs) who are least likely to have access to quality healthcare have the highest proportion of those living with HIV. Mobile technology has played an important role in providing access to information and services among adolescents within the region in recent years. This review aims to synthesise and summarise information that will be useful in planning, designing, and implementing future mHealth strategies within the region.

**Methods and Analysis:** Interventional studies on the prevention and management of HIV among adolescents that used mobile technology in LMICs will be included. MEDLINE (via PubMed), EMBASE, Web of Science, CINAHL, and the Cochrane Library are the information sources that have been identified as relevant to the area of study. These sources will be searched from inception to March 2023. The risk of bias will be assessed using the Cochrane Risk of Bias tool. The scalability of each study will be assessed using the Intervention Scalability Assessment Tool (ISAT). Two independent reviewers will conduct the selection of studies, data extraction, assessment of the risk of bias, and scalability. A narrative synthesis of all the included studies will be provided through a table.

**Ethics and dissemination:** An ethical approval was not necessary for this study. This is a systematic review of publicly available information and therefore ethical approval was not deemed necessary. The results of this review will be published in a peer reviewed journal and dataset will be presented in the main manuscript.

**Strengths and limitations:**

- We believe that the likelihood of missing any published article will be low because of the information sources we are considering.
- The scalability tool (ISAT) has not been used in any systematic review before.
- The evidence provided in this review will be limited to low-middle-income countries.
- The exclusion of studies not published in English is a limitation for this review.

## Introduction

In recent times, there has been an increase in the global ownership and use of mobile phones, with about 5.6 billion unique users of mobile phones [1]. The global increase in the use of mobile phone can be primarily attributable to its rapid adoption in developing countries [2]. For example, an increase in the ownership of smart phones in LMICs such as Ghana, Senegal, Philippines, Jordan, and Lebanon were reported between 2015 and 2017. In Lebanon, an increase in smart phone ownership from 52% to 80% between 2015 and 2017 was reported [3]. In Nigeria, the Nigerian Communications Commission (NCC) also reported that the total number of cellular network subscribers has increased by 9.4% between May 2021 and May 2022 [4]. Further, a report has shown that there has been an increase in the number of women in LMICs who have access to mobile internet in 2022 compared to the 2017 data[5]. The increased access to mobile phones (especially smartphones) has facilitated their use in different aspects of life and livelihood, including healthcare.

The World Health Organization (WHO) acknowledges that leveraging on advancements in technology is a way to optimize service delivery, and these advancements and innovations in technology offers new ways to provide quality services at affordable costs[6,7]. Advantages of the use of mobile technologies in health care include improvement in the speed and efficiency of diagnosis and initiation of treatment, remote evaluation of patients, and increased access to risk-reduction education and knowledge of different health issues. Despite the numerous advantages and the proliferation of use of mobile phone technology in healthcare, some challenges have been recorded around issues of usability of interventions, mobile network coverage, ethics, data security and privacy, reliability of intervention, proficiency and integration of technologies used[8].

However, the increasing use and dexterity of mobile phone users have enhanced the application of mobile phones to address public health problems [9–11]. The increasing development in the usage of mobile phone technology among young people provides more modalities to satisfy their health demands [12,13]. Mobile phones have been used to improve young people’s sexual and reproductive health [14], physical health [15], and HIV prevention and management [11]. However, some researchers are worried that the use of mobile phones in health interventions may further heighten the disproportionate access to health information and services as only individuals with access to mobile phones, internet services and required knowledge and skills to operate a smartphone will be able to utilise such services [16]. Nevertheless, the use of mobile phones in providing health information and services is greatly on the increase, especially among adolescents and young people [12].

HIV/AIDS remains a global health issue and is one of the first five leading causes of death among adolescents in sub-Saharan Africa (ref). However, adolescents are disproportionately affected by HIV/AIDS. Reports show that adolescents account for about 5% of people living with HIV and 11% of all new infections globally[17]. Most of these adolescents (89%) live in developing countries with poor access to adequate healthcare[17]. Several studies have been conducted to prevent and manage HIV among this age group. Mobile phones have played significant roles in planning and executing HIV programmatic objectives [9].

Several reviews have been conducted on the use of mHealth tools in support of HIV programmatic priorities [9–11]. These reviews focused on low- and middle-income countries (LMICs), including countries in Africa. All the reviews reported increased use of mobile phones for intervention within the regions. One of these reviews reported that text messaging was the main mechanism of delivering mHealth interventions in Africa [10]. However, a recent review reported that mobile applications and web-based mobile interventions are now more commonly used in mHealth interventions[11].

Besides showing the increase in the use of mobile phones in health interventions, previous reviews have also shown that mobile interventions are effective in improving some health outcomes among young people [12] and the general population [18,19]. A review reported the effectiveness of mHealth across various health outcomes [19]. In HIV research, mHealth interventions have been shown to improve knowledge of HIV among young people and reduce the practice of some health risk behaviours [11]. It has also been shown to increase medication adherence resulting in improved quality of life among adolescents [19].

Although several studies have shown that mHealth interventions on HIV/AIDS among adolescents are effective, acceptable, feasible, and cost-effective[11], very few have reported on the scalability of mHealth interventions. The increased use of mobile phones in the prevention and management of HIV because of increased access to mobile phones among young people provides a huge opportunity and potential for most interventions to be scaled up to a larger population.

A recent review evaluated the acceptability, adoption, appropriateness, cost, feasibility, fidelity, penetration, and sustainability of mobile phone interventions for HIV prevention among young people in LMIC [11]. This review reported the different delivery modalities such as text messages, mobile applications, and web-based mHealth interventions and the components of mobile interventions in LMICs. However, it only focused on reporting HIV prevention outcomes such as education, SRH counselling, and self-testing.

This proposed review seeks to assess the scalability of different mHealth interventions on HIV among adolescents, provide evidence on the potential of mobile technology in HIV prevention and management, and identify good practices from studies in the development of mHealth programmes. Although there has been increase in the use of mobile technology for health services, there have been limited empirical evidence on assessing the scalability of these interventions[20]. This can be mostly attributed to the fact that most intervention research do not plan for scalability from the design phase of the intervention. The lack of plan for scale-up of health interventions has been identified as a barrier for scalability[21]. Other barriers include lack of technical consensus and health inequities in accessing health care[22,23]. Inequity in access to health care is a common problem for prevention and management of HIV among adolescents, especially in SSA. Adolescents and young people are least likely to have access SRH/HIV information and management services. Reports have shown that adolescents are more likely to use mobile technology in search of information. It is therefore assumed that the designing and implementing interventions using mobile technology will increase access to information and services among this age group. Some of these interventions have been shown to be successful in improving access to information, adherence, and testing[11], therefore a scale-up of these effective interventions may improve prevention and management of HIV among adolescents.

We define scalability in line with Milat *et al*., 2013 as ‘the ability of a health intervention shown to be efficacious on a small scale or under controlled conditions to be expanded under real-world conditions to reach a greater proportion of the eligible population, while retaining effectiveness’[24]. In our review, we intend to build on existing evidence and consider how mHealth facilitates HIV prevention and management among adolescents in LMICs. Our review will consider all HIV prevention and management strategies including outreach and education, SRH counselling, HIV testing and counselling, linkage to care, CD4 screening, ART initiation, and treatment adherence among adolescents and how mHealth interventions have facilitated these strategies. We will assess scalability of these interventions using a scoring tool that assesses various domains related to scalability such as strategic content, evidence of effectiveness, program cost, fidelity, and adaptation, reach and acceptability, delivery setting and workforce, implementation infrastructure and sustainability.

## Methods and analysis

### Research objectives

The primary objective of this review is to assess the scalability of mobile technology-based interventions in the prevention and management of HIV/AIDS among adolescents in low- and middle-income countries. Other objectives include Identify and understand the modality of the use of mobile technology-based interventions in the prevention and management of HIV/AIDS among adolescents in low- and middle-income countries.

Report evidence of effectiveness of mobile technology in HIV prevention (including testing, counselling, and knowledge of HIV) and management (including adherence, support, and retention in treatment), and Identify good practices from studies reviewed in the development of mHealth programmes.

### Eligibility criteria

#### Inclusion criteria

1. We will include randomised controlled trials and non-randomised controlled trials. Studies using quasi experimental methods with comparison groups, but no random assignment will also be included in the review.
2. We will include published peer-reviewed articles as well as unpublished literatures such as project reports and ongoing studies where preliminary findings are available to us [25].
3. Studies conducted in low- and-middle-income countries as defined by the World Bank[26].
4. Studies involving adolescents (boys and girls) aged 10 to 19. Studies conducted among young people including individuals older than 19 years will also be included in the review if adolescents (10-19 years) are included in the study.
5. Studies that examined the impacts of the use of mobile technology on HIV prevention and management among adolescents.
6. The comparison (control) group in each included study can be participants who did not use mobile technology or any other interventions, or participants who received alternative interventions.
7. No restrictions will be placed on the year of publication, and sample size of the study, or the duration of the intervention.

#### Exclusion criteria

All studies that do not fulfil the criteria listed above will be excluded from this review. Examples of such studies include

1. Studies not reporting primary data. Non-original research, secondary reports, commentaries, editorials, and reviews will be excluded from this review. We will only include studies with original data; therefore, nonempirical studies such as letters, perspectives, and editorials will not be included.
2. Experimental studies that did not account for the baseline differences between intervention arms will not be included
3. Observational studies such as cross-sectional studies
4. Studies that were not published in English language.
5. Studies on use of mobile technology in HIV prevention and management conducted only among adults older than 19 years will be excluded.
6. Studies that described mobile technology usage only without linkage to specific prevention and management of HIV among adolescents.

### Information sources

A systematic search will be conducted to identify eligible peer-reviewed literature in the following databases: from the inception of each database through March 2023: MEDLINE (via PubMed), EMBASE, Web of Science, CINAHL, and the Cochrane Library. Reference lists of included papers were manually searched for additional relevant citations. We will also search ClinicalTrial.gov and WHO International Clinical Trials Registry Platform. These electronic databases were selected based on consultations and a brief review of relevant reviews conducted in the past. Additionally, ‘cited by’ tool in Google Scholar was used to identify potentially relevant studies. We searched other governmental or organisational websites, such as World Health Organisation, United Nations ICEF, UNFPA and World Bank for studies or ongoing studies with preliminary results not identified from the database searching. We will also screen and search the references of the related recent systematic reviews. Further, if we require more information to confirm eligibility of a study, we will contact the authors by mail. Authors will be mailed a maximum of two times. After the initial mail, a reminder will be sent after one week if there was no response to the initial mail. After the second mail, we will wait a maximum of one more week after which we will proceed to exclude the study.

### Search strategy

The search strategy was developed in consultation with a health science librarian at the University of Ibadan and senior colleagues who have conducted similar reviews. Previous reviews on the similar topic were also consulted in developing the search strategy. The PubMed search builder was used. We created search terms using a combination of Medical Subject Headings, keywords, and phrases, including “HIV” or “prevention” in combination with, but not limited to, any of the following: “eHealth”, “mHealth”, “smartphone”, “mobile phone”, “mobile application”, “app”, “internet”, “technology”, and “adolescence” or “adolescents” or “young people”.

The PubMed search strategy was developed as the primary search strategy template and adapted for the other databases. The initial search was carried out between December 2021 to February 2022. A repeat search was conducted in June 2022 and December 2022. An updated search will be conducted in March 2023. The final search strategy used is provided in supplementary table 1.

### Data management and selection of studies

Mendeley (Elsevier) will be used to store the articles retrieved from the electronic databases. Mendeley functions will also be used to identify and delete duplicate records. A manual duplication check will also be conducted after the initial check on Mendeley is done. Removal of duplicates will be conducted prior to screening.

Studies will be screened in two stages. In the first stage, titles and abstracts will be screened to exclude ineligible studies, using a broad and customised checklist for study selection. Full-text versions of selected abstracts will then be downloaded/retrieved and assessed independently by the two reviewers to ensure that inclusion criteria are met. The two independent reviewers will be researchers who are familiar with the concept of the review and have knowledge of the selection criteria. Screening and selection of studies will be facilitated by the creation of appropriately labelled subfolders in Mendeley, to segregate studies for inclusion and exclusion. Specific reasons for study exclusion will be documented and reported using the Preferred Reporting Items for Systematic Reviews and Meta-Analyses (PRISMA) flow diagram [27]. The results of each step will be compared, and inconsistencies or conflicts will be resolved through discussion or arbitration from a senior colleague. The reviewers will not be blinded to the details (such as journal name, names of authors of the articles).

Scalability of the studies identified in this review will be conducted using the Intervention Scalability Assessment Tool (ISAT)^1^. The ISAT is a tool that facilitates assessment and decision making on the potential scalability of population health interventions and demonstrates its potential for use in a real-world setting. The tool is divided into three main parts. The first two parts (presented in Table 1) contain five domains covering aspects of the scale-up context and proposed implementation requirements [28]. The third part is a summative assessment on the scoring from the first two parts to generate a radar plot against which the readiness for scale-up can be assessed (Table 2 describes these scores) [29]. We will also contact authors for information in completing the scalability assessment tool. We will follow the same procedure for contacting the authors described above in information sources.

**Table 1.** Intervention Scalability Assessment Tool (ISAT) domains and objectives

| Domain | Objective of the domain |
| --- | --- |
| Part A |  |
| A1: The problem | Consideration of the problem that is being addressed. The questions in this domain seek a description of the problem, who it affects, what it affects and how it is currently being addressed (if at all). |
| A2: The intervention | Description of the proposed program/intervention to address the problem. |
| A3: Strategic/political context | Consideration of the current strategic/political/environmental contextual factors that are potentially important influences on any intervention to be scaled up. |
| A4: Evidence of effectiveness | Consideration of the level of evidence available to support the scale-up of the proposed intervention, such as scientific literature and/or other known evaluations of the intervention. |
| A5: Intervention costs and benefits | Consideration of the known costs of the intervention delivery as well as any quantifiable benefits. This includes the results of any types of economic evaluation studies. |
| Part B |  |
| B1: Fidelity and adaptation | Consideration of whether there are any proposed changes to the intervention required for scale-up. |
| B2: Reach and acceptability | Consideration of the reach and acceptability of the intervention for the target population. |
| B3: Delivery setting and workforce | Consideration of the setting within which the intervention is delivered as well as the delivery workforce. |
| B4: Implementation infrastructure | Consideration of the potential implementation infrastructure required for scale-up. |
| B5: Sustainability | Consideration of the potential longer-term outcomes of the scale-up and how, once scaled up, the intervention could become sustainable over the medium to longer term. |

**Table 2:**
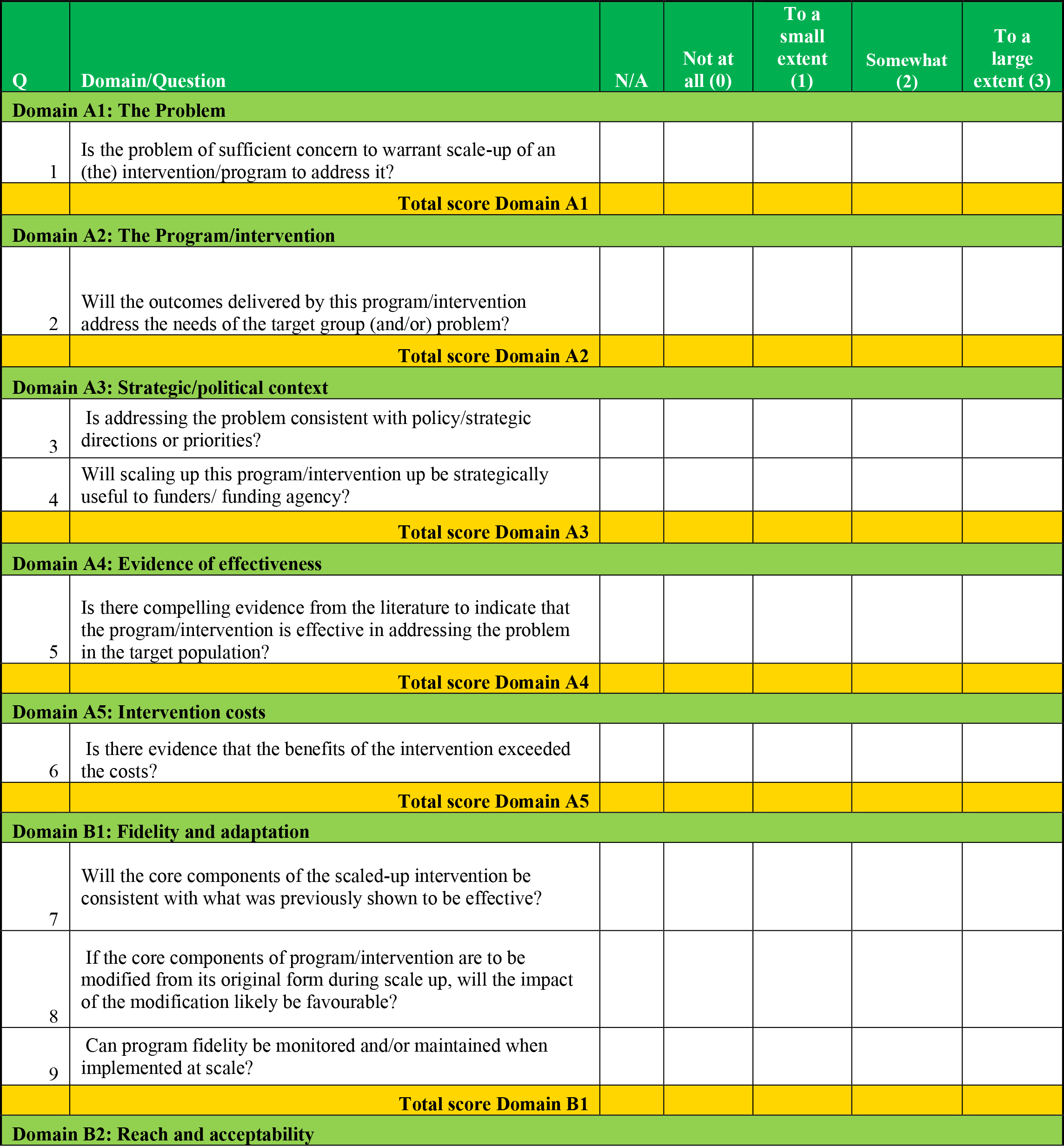
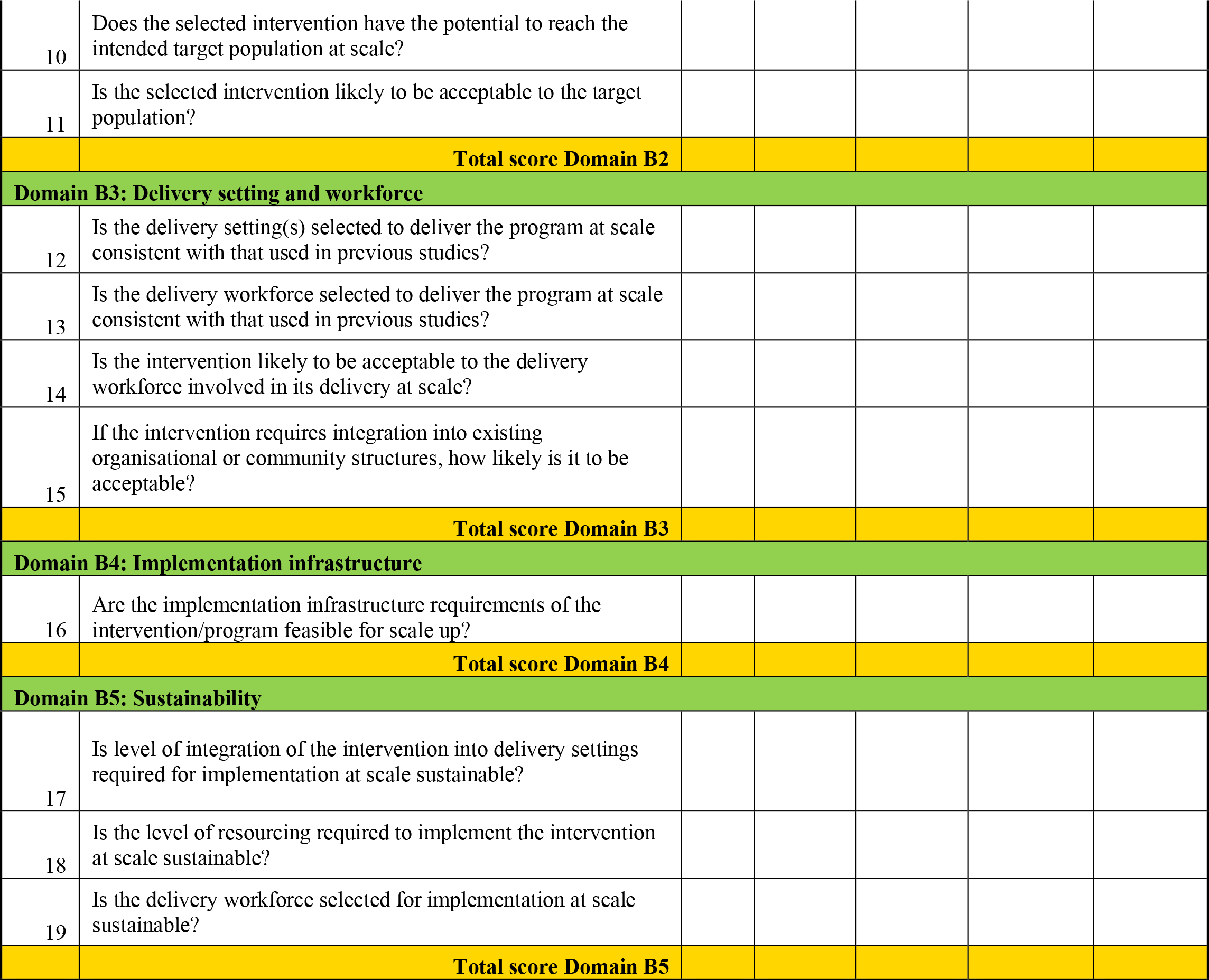
The ISAT score sheet

### Data extraction

Data from the full text of selected studies will be extracted by two independent reviewers (the same reviewers involved in the selection of the studies to be included in the reviews). A data extraction form (Table 3) will be used in the extraction of the study. This data extraction form was designed using the PICO framework. We will extract information about the population of interest (including age, gender, and schooling status), method and content of intervention, and the outcome of interest. The extraction form will be pretested with at least five randomly selected studies. If there are disagreements in the extracted information, differences will be resolved through discussion or by a senior colleague.

**Table 3:** Data extraction sheet

| Title | Author | Journal or source | Year of publication | Study location | Study design | Recruitment method and Sample size | Target Sample characteristics |  |  | Timing of intervention | Method and content of intervention |  |  |  |  | Outcomes | Main findings | Scalability score | Scalability recommendation |
| --- | --- | --- | --- | --- | --- | --- | --- | --- | --- | --- | --- | --- | --- | --- | --- | --- | --- | --- | --- |
|  |  |  |  |  |  |  | Age group (early, mid, and late adolescence) | Sex | Schooling status |  | SMS based | web based | Mobile app | Call | Intervention component |  |  |  |  |

### Assessment of risk bias

To assess risk of bias, we will use the version 2 of the Cochrane Risk of Bias tool [30]. The tool considers the following domains of bias: Bias arising from the randomisation process, bias due to deviations from intended interventions, bias due to missing outcome data, bias in measurement of the outcome, and bias in selection of the reported result. Authors of the selected studies will be contacted in completing the risk of bias tool. We will follow the same procedure described in information sources above for contacting authors. All selected studies will be scored as low, some concerns and high risk of bias. To assess risk of bias in non-randomised studies of interventions we will use the ROBINS-I tool. The tool considers seven domains of bias: bias due to confounding, selection of participants, classification of intervention, deviation from intended intervention, missing data, measurement of outcomes, and selection of reported results. Studies will be scored across domains and reported as having low, moderate, serious, or critical risk of bias. We will also contact authors of these studies for further information to complete the ROBINS-I tool.

### Registration and reporting

This protocol has been submitted for registration with the International Prospective Register of Systematic Reviews (PROSPERO). Registration number is CRD42022362130.

### Data Synthesis and Interpretation

The main characteristics and key findings from the selected manuscripts will be summarised in a table as shown in table 2. This table used in data extraction will be a major analytical tool. From the table, the details of findings to provide responses for the three objectives will be summarised. For the first objective, the modality of the use of mobile technology in prevention and management of HIV among adolescents, the methods and context of use of mobile technology and the intervention components will be analysed to present the different modalities of the use of mobile technology in preventing and managing HIV among adolescents. Information for the second objective will be provided through the summary of outcomes from each identified study. The evidence of effectiveness as reported by the selected studies and the reach and acceptability of the intervention among the target population will be summarised and presented. To identify good practices, the study methods including the study design, retention activities, and recruitment methods will be summarised and presented in line with positive outcomes for best practices. The fidelity and adaptation, that is, consideration of whether there are any proposed changes to intervention required for scale-up, presented, or suggested by the authors will also be summarised and presented as best practices. We will compare these findings in high-quality studies and low-quality studies. The practices of the high-quality studies with effective findings will be reported as best practices.

Risk of bias will be determined and scored by two independent reviewers. The scores for individual manuscripts will determine the overall risk of bias od the body of evidence in this systematic review. The final scalability score will be determined through the addition of the scores across the five domains of the ISAT tool. Scores for each domain will be imputed into the ISAT scoring sheet in excel and a radar plot will be generated. The radar plot will enable a visual comparison across the domains. For each domain, the scores range from a minimum of 0 to a maximum score of 3. The summary scalability score will be an average of all the domain scores, with a minimum and maximum obtainable score of 0 and 3 respectively. Each manuscript will be initially considered by a group of reviewers. This group will include intervention and policy researchers. Prior to the group meeting, authors will be contacted to obtain other information necessary to fill the ISAT. For example, information about if the study was considered for scale-up. A discussion to determine the appropriate score to assign per domain per manuscript will be held. A recommendation will be made per manuscript. The recommendations will include merit scale up (score of ≥2), promising, but further information/planning is warranted (<2≥1) and does not merit scale up (<1). We will document all processes involved in the assessment for scalability and publish this for transparency on how the scalability scores were arrived, including manuscripts reviewed to aid in the assessment.

### Patient and Public involvement statement

No patient involved in this study

### Ethics and dissemination

We did not seek for ethical approval for this study because all data that will be used in this study are publicly available. The results of this study will be disseminated through presentation at a scientific conference and publication in a peer-reviewed journal. The dataset will be published as part of the main manuscript.

## Discussion

There is a problem of health underdevelopment which leads to adverse outcomes among adolescents and young people who are already vulnerable, especially adolescents and young people living in SSA. Solving the problem of health development requires a holistic approach which the introduction of mHealth has the potential to bridge. The use of mHealth has the potential to promote uptake as well as improve the availability of health information and services to everyone including underserved and vulnerable populations, thereby narrowing the gap, and promoting universal health coverage.

Adolescents in SSA make up a large percentage of adolescents globally. Unequal access to health information and services within the region subjects most adolescents globally to poor access to health care. Although adolescent sexual and reproductive health is one of the most funded domains of adolescent health, adolescents in the SSA region still lack access to comprehensive health information and services. About 11% of new HIV infections occur among adolescents and more than 80% of these adolescents live in SSA[31]. It is important to design and implement programs that promote the prevention and management of HIV to reduce the rate of new infections and mitigate the effect of HIV among adolescents. Programs which utilize the mHealth intervention have the potential to promote adherence to drug regimen and provide adolescents with useful information which will promote their health and wellbeing.

Although adolescents are vulnerable and largely affected by HIV, cultural, religious, and political sensitivities influence adolescents’ access to adequate information and services[32–34]. Designing programs with the help of mHealth can provide safe platforms to access information and services to prevent HIV infection. It will also help promote safer sexual behaviour and improve adherence to the management of HIV among positive ones, thereby improving their quality of life. Also, considering that adolescents make up almost a third of the world’s population, addressing HIV prevention and management among this age group will remarkably affect the nations’ development positively.

Research provides us with useful and evidence-based information that helps us make informed policies and decisions regarding what works and what does not. However, it has also been shown that some interventions that are effective at a small scale are not so effective on a large scale, and some interventions that are effective in a controlled study may not be so effective in the real world. Therefore, planning for scale up at onset and measuring the scalability of interventions becomes an essential aspect of planning, designing, and implementing an intervention program[20]. In this review, we intend to report how scalable mHealth interventions are within LMICs. This review will provide details of good practices that make some studies more likely to be successful at scale than others. Although there are several scale-up frameworks, for this review we chose the Milat et. al., 2013; 2019; 2020 framework because it emphasises evidence effectiveness as a precondition for scale-up [20,24,29]. However, we recognise that this tool has never been used in a systematic review nor by researchers to assess scalability of interventions. Therefore, this poses a potential limitation. Further, although we will be mailing authors and obtaining information to complete the ISAT, we recognise that we may not hear back from all authors and that not all studies might have been considered for scale-up. However, adapting this scalability assessment tool, provides feedback for the developers and an opportunity to improve on the tool for future assessments.

Conclusively, adolescents and young people in LMICs are disproportionately affected by HIV infection and are also less likely to have access to appropriate health information and services. Some of the interventions using mobile technology have shown great promise in bridging this gap. However, beyond providing information on the effectiveness of such interventions, it is critical to be able to prepare and plan for the application of such interventions at scale. This paper describes the protocol we intend to employ in reviewing studies that used mobile technology in the prevention and or management of HIV among adolescents 10-19years.

## Data Availability

All data produced in the present study are available upon reasonable request to the authors

## Authors’ contributions

EA conceptualized the study with significant input from DQ, AOO, AO and WF. EA wrote the first draft of the paper with input and revisions from DQ, AOO, AO and WF. All authors read and approved the final version of the paper.

## Footnotes

1 Authors have been contacted and permission to use the tool has been granted

